# A Genomics England haplotype reference panel and the imputation of the UK Biobank

**DOI:** 10.1101/2023.11.06.23298035

**Authors:** Sinan Shi, Simone Rubinacci, Sile Hu, Loukas Moutsianas, Alex Stuckey, Anna C Need, The Genomics England Research Consortium, Mark Caulfield, Jonathan Marchini, Simon Myers

**Affiliations:** Department of Statistics, University of Oxford, Oxford, United Kingdom; Harvard medical school, Harvard University, Boston, United States; Novo Nordisk Research Centre, Oxford, United Kingdom; Genomics England, London, United Kingdom; Queen Mary University of London, London, United Kingdom; Regeneron Genetic Center, Tarrytown, New York, United States

## Abstract

The choice of reference panels significantly impacts phasing, imputation and GWAS results. In this study, we built a haplotype reference panel using the Genomics England (GEL) high-coverage sequencing dataset, one of the largest genetic variation resources ever collected in the UK. The resulting reference panel consists of 156,390 haplotypes and 342 million autosomal variants. The GEL reference panel demonstrates reliable imputation of variants as rare as 1 in 10,000 within the White British population, with an imputation r^2^ value of 0.75. The resulting imputed UKB data (GEL-UKB) contains three times more variants, predominantly rare variants, compared to the UKB data previously imputed using the HRC and UK10K reference panel. The GEL-UKB presents a unique opportunity for the reliable discovery of rare associations across the whole genome, especially within the regions not covered by the exome sequencing data. Rare variant signals with high confidence are predominantly from rare coding variants, implying firstly, a probable tendency for existing rare non-coding mutations to not reach a disruptive level comparable to that of coding variants. Secondly, it raises the possibility that the current sample size of UK Biobank may be insufficient for detecting rare variants with a moderate effect size, even with the whole genome sequencing. The resulting GEL phased haplotype reference panel has been made available on the GEL platform and widely used by GEL users. Our GEL imputed UKB data has been adopted as one of the UKB official imputed data resources (Data Field 21008).

## Main

Genomics England (GEL) has carried out whole genome sequencing (WGS) of over 120,000 genomes from over 80,000 individuals taking part in the 100,000 Genomes Project, using an average sequencing coverage depth of ∼30x^1^. The recruitment strategy focussed on patients with rare disease (disorders affecting < 1 in 2000 people) and cancer, and their close relatives, across hospitals in England. We constructed a GEL phased reference panel based on 78,195 high-coverage sequencing germline genomes, with a diverse ethnic representation. The high degree of relatedness among the samples enhances the power of filters, such as the Mendel error filter, for eliminating false positive variant sites identified in the sequencing data, and also leads to more accurate phasing and imputation of rare variants. In particular, it enables even variants found in only one or two individuals to be phased through transmission, a task which is more difficult in the absence of related samples or phase information in sequencing reads^2^.

The resulting GEL reference panel consists of 341,922,205 autosomal variants, with 31,502,703 (9.26%) being INDELs with an average length of 5bp and a maximum length of 50bp. The majority of the variants in the GEL reference panel are rare. 287.2 million (84.1%) of identified variants possess an allele frequency lower than 0.0001, including 66.7 million (19.5%) singletons and 91.1 million (26.7%) doubletons. We compared the variants in GEL reference panel to the widely used TOPMed r2^3^ and HRC^4^ panels and found GEL has 8 times and 1.1 times more variants than the HRC and TOPMed panels respectively (**Figure 1b** and **Supplementary Figure 1**). Due to the use of mostly low coverage sequencing technology, the HRC dataset has limited numbers of rare variants, especially those with AF ≤ 10^−4^. While the numbers of rare variants captured in TOPMed and GEL are similar, around half of the ultra-rare variants (AF ≤ 10^−4^) from GEL and TOPMed are non-shared across the panels (**Supplementary Figure 1**). As expected, all three panels capture a similar set of more common (AF>10^−2^) variants, with less than 4% unique to each panel (**Supplementary Figure 1**), indicating common variants are largely saturated.

**Figure 1:**
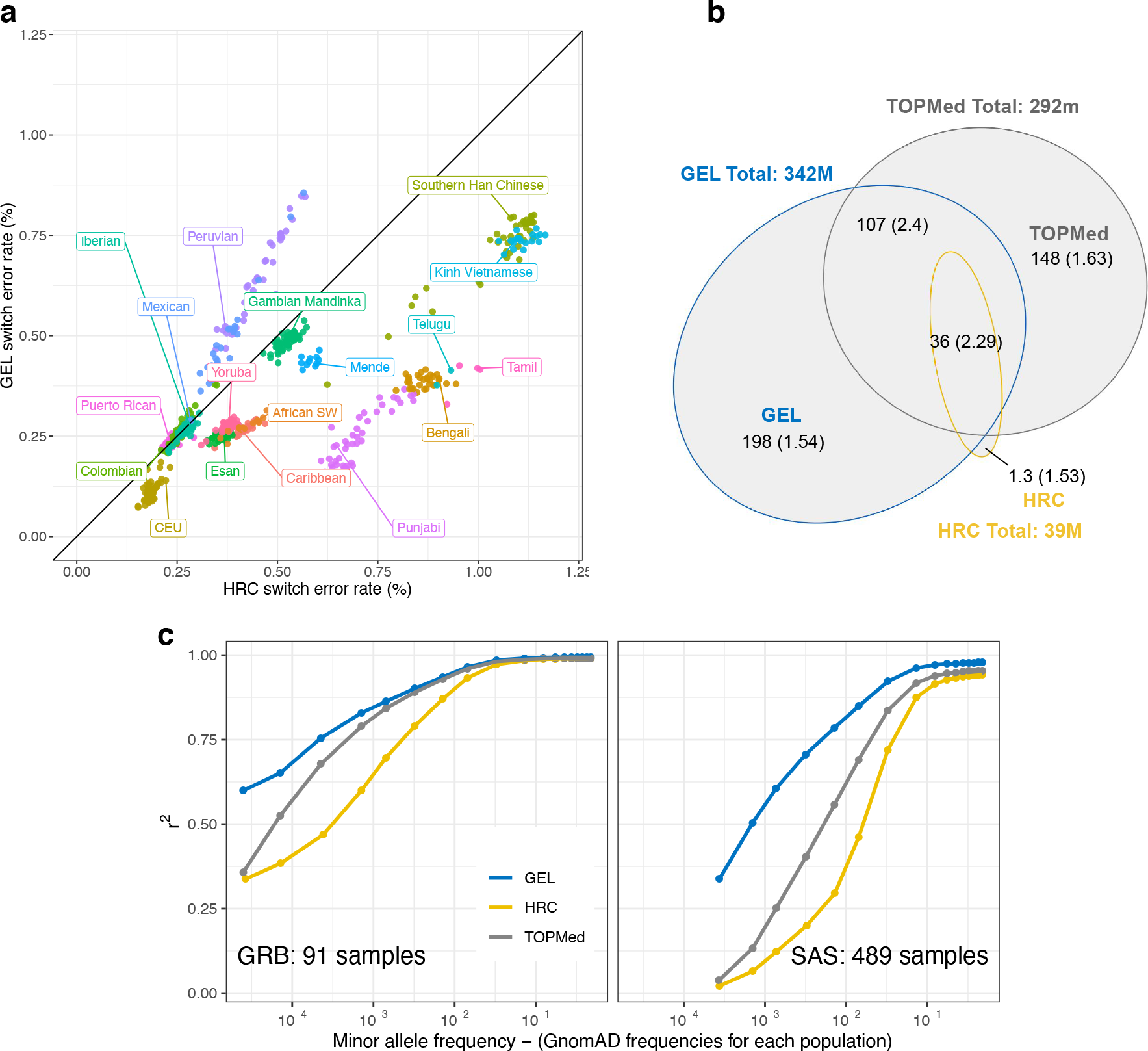
a) Phasing quality for 589 high coverage 1,000 Genome children from mother-father-child trio families, using HRC and GEL reference panels. b) Venn diagram comparing numbers of variants from the GEL, HRC and TOPMed reference panels. The numbers show the variant count (in millions of variants), followed by the Ts/Tv ratio of these variants in brackets. c) Imputation performance, measured by r^2^ (**Methods**), for imputation of 1000 genomes samples from the *White British* (left) and South Asian (right) groups, using three different reference panels (labels). The variants are stratified by GnomAD allele frequency (v3.3.1)^6^ of their corresponding population.

The GEL reference panel can be used as a powerful resource for phasing European and South Asian samples, due to their strong representation in the dataset. We compared the phasing accuracy achievable using the GEL and HRC reference panel across 26 diverse populations from the 1000 Genomes project (**Methods**). GEL phasing of these samples achieved lower switch error rates than HRC phasing, across the CEU (Northern European from Utah), African, South Asian and East Asian ancestry populations (**Figure 1a)**, with HRC only showing improved performance for South American samples, which are not significantly represented in GEL. GEL phasing switch error rates are 0.18%, 0.33%, 0.31% and 0.73% for European, African, South Asian and East Asian samples respectively.

A primary use of the GEL will be as a reference panel for genotype imputation of other datasets. We assessed the imputation accuracy among 2,405 1,000 Genomes samples, using the GEL, TOPMed and HRC reference panels. We used genotypes at the 716,473 autosomal bi-allelic SNP positions on the UK Biobank Axiom array^5^ to impute all non-array sites using each reference panel (**Methods**). Squared correlation *r*^2^ between the imputed allele dosages and true genotypes were calculated, stratified by the independently estimated gnomAD (v3.3.1) minor allele frequency^6^. As we focus on showing the overall performance of the reference panel across different allele frequencies, only variants present within gnomAD are shown. As a result, the number of tested variants differs across reference panels. GEL achieved higher *r*^2^than HRC in all allele frequency bins for all ethnicities (**Supplementary Figure 4)** and outperforms the TOPMed panel in White British (GBR) and South Asian (SAS) samples, especially for rarer variants: at MAF < 10^−5^, the GEL imputation *r*^2^ for GBR samples is 0.6, compared to 0.3 and 0.29 using TOPMed and HRC, respectively (**Figure 1c**). The TOPMed panel outperforms GEL in African, American and East Asian samples due to its better representation from these groups (**Supplementary Figure 4**).

We used the GEL panel to impute 488,315 UK Biobank samples at 342,573,817 variants, producing a “GEL-UKB” dataset; we compared to the corresponding HRC and UK10K-imputed “HRC-UKB”^5^. GEL-UKB has around 3 times more variants than HRC-UKB, 3.5 times more missense variants, and 6.6 times more “high impact consequence” variants (**Supplementary Table 5**). The imputed information scores (**Method**) were higher for GEL-UKB than HRC-UKB for 87% of the variants that are in common, while 98% (78%) of GEL-imputed variants at frequency below 10^−4^ (10^−5^) exceeded a threshold of 0.3, vs 78% (54%) for HRC (**Supplementary Figure 2-3**).

**Figure 2:**
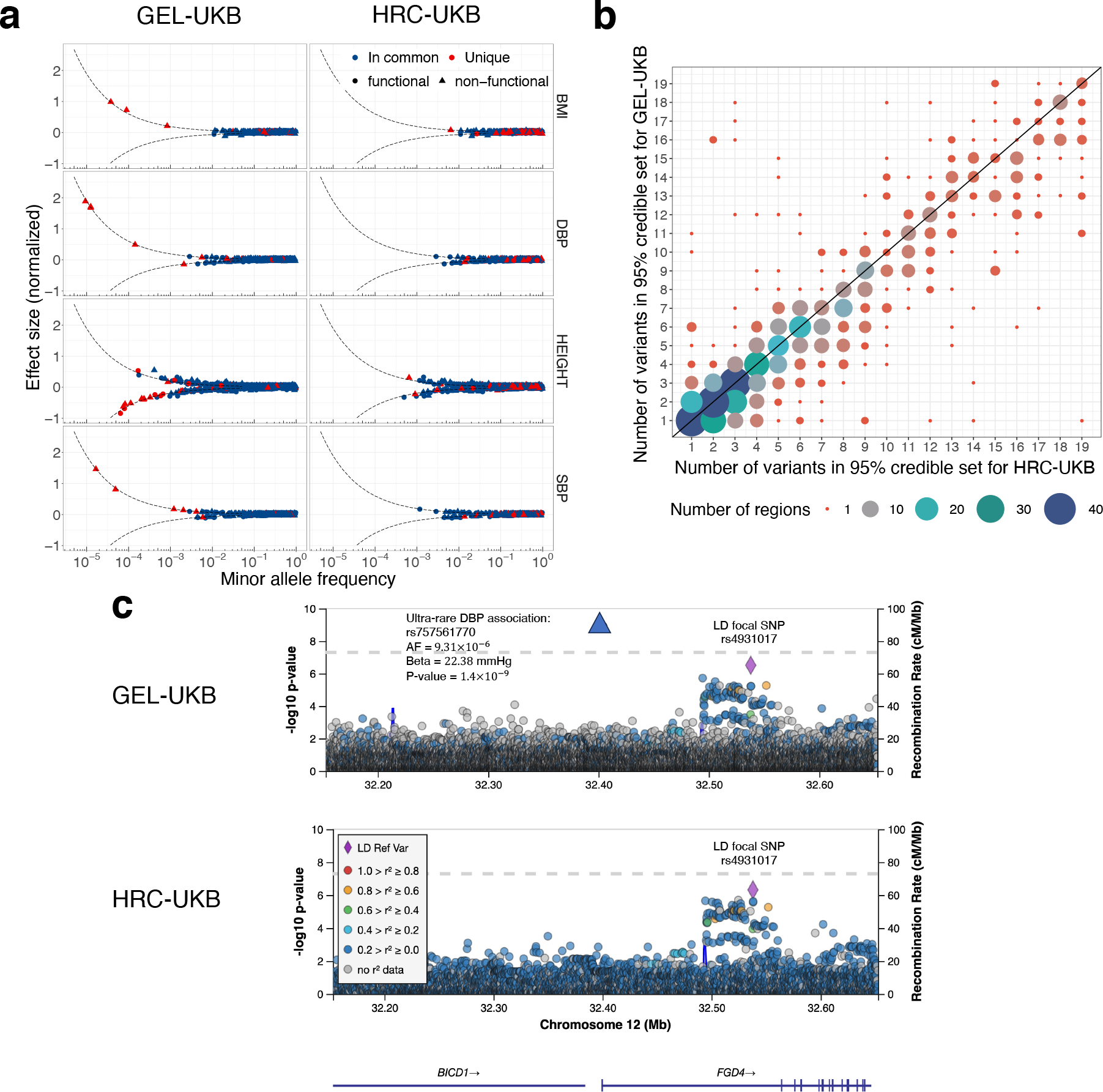
a) A set of independent genome-wide significant (*p* < 5 × 10^−4^) associations identified by step-wise regressions (conditioned joint analysis), and with INFO > 0.8, are plotted versus their imputed allele frequency (x-axis). The blue colour represent variants that were flagged by step-wise regressions in one dataset and also showed a significant GWAS association in the other dataset; The red colour indicates that the variant is unique to each dataset. The shape of the data points reflects the predicted consequences of the variants as determined by VEP. Dots represent functional variants, including stop gained, stop lost, splice donor/acceptor, frameshift, in-frame insertion/deletion, and missense and the triangles indicate non-functional variants. The dotted lines indicates the smallest effect sizes that can be captured by the p-value threshold (*p* < 5 × 10^−4^). b) Comparison of the number of variants in the 95% credible sets for GEL-UKB and HRC-UKB fine-mapping results for standing height (capped at 20 variants; **Methods**). The circle sizes represent the number of fine-mapping regions showing each combination; plots below the diagonal correspond to GEL-UKB having fewer variants in the credible set compared to HRC-UKB. c) The LocusZoom plot of ultra-rare variant association (rs757561770) detected by GEL-UKB. The color indicates the LD between SNPs and the focal SNP rs4931017, showing that rs757561770 is in low LD with the focal SNP (*r*^2^ = 6.57 × 10^−5^). The blue lines show the recombination rate of the region.

To demonstrate the use of GEL-UKB, exemplar GWAS were carried out on four quantitative traits, including standing height (HEIGHT), body mass index (BMI), systolic (SBP) and diastolic (DBP) blood pressure, with variant testing using REGENIE^7^. Across all four traits, we found 31,699 and 30,711 significant (P-value < 5 × 10^−8^) rarer variant associations (MAF < 0.05) from GEL-UKB and HRC-UKB, respectively. The GEL-UKB common variant associations were also less likely to be subjected to false associations than HRC-UKB (**Supplementary Notes; Supplementary Table 2; Supplementary Figure 6-8**). A recent exome-sequencing based association study reported 31, 0, 1, and 2 rarer (MAF < 0.05) genome-wide significant (P-value < 2.18 × 10^−11^) variant-trait associations across HEIGHT, BMI, SBP and DBP, respectively^8^. We discovered 70% of these associations using GEL-UKB, compared to 56% using HRC-UKB at the same p-value threshold. Relaxing the GEL p-value threshold to 5 × 10^−8^, GEL-UKB identifies 76% of these associations (**Supplementary Table 3**). When we compare to the UKB whole exome imputation results^9^, all but 4 out of the 28 exome imputation likely-causal rare coding variants associated with standing height(p-value < 5 × 10^−8^) are found to be significant using GEL-UKB, whereas all but 9 of such variants are found to be significant using HRC-UKB (**Supplementary Figure 9**).

This comparison in the exonic portion of the genome provides confidence that whole-genome imputation using GEL can identify most associations directly observed using sequencing. We next compared the performance of GEL-UKB to the widely used imputed genotypes available for the full set of UKB samples, HRC-UKB, and we examined those novel associations identified using GEL-UKB. First examining shared signals (mainly at common sites), we saw a useful improvement in fine-mapping (**method**) using GEL-UKB vs. HRC-UKB. 44% of the GEL-UKB based 95% credible sets contain fewer SNPs, while 25% contain more SNPs (**Figure 2b; Supplementary Table 4**), with the remainder identical in size.

A more dramatic difference is observed for rare variants: independent rare variant associations (MAF < 5 × 10^−4^), accompanied by high estimated effect sizes (**Figure 2a)** required to reach statistical significance at these frequencies, are almost exclusively discovered by GEL-UKB (**Figure 2a)**. For example, GEL-UKB detected a novel ultra-rare association signal for DBP at rs757561770 in FGD4, with allele frequency 9.31 × 10^−6^. Common variants in FGD4 has previously been reported to be associated with hypertension^10^ (**Figure 2c**). Interestingly, this SNP is intronic and does not show strong linkage disequilibrium (r^2^ > 0.7) with any coding variant within the GEL panel **(Supplementary Table 6**).

Because we test the entire genome, our results allow us to investigate whether large-effect mutations (which in our example GWAS are only found at low frequency; **Figure 2b**) occur in coding or non-coding DNA. We identified 27 independent large-effect/rare-variant signals (AF < 0.001), across the four traits using step-wise regression (**Method**). Of these, 17 were either coding (n=7) or in strong LD (r^2^ > 0.7) with a coding variant. An additional 1 is associated with splice-site variants and 2 with variants in 5’ UTRs or 3’ UTRs of genes (**Supplementary Table 6**). In total, 62% of all the rare variant associations and 88% of the strongest associations (p-value <2.18 × 10^−11^) were associated with genic sequences (**Supplementary Table 6**). If replicated for other phenotypes, this implies that it could be likely rare for variation in other non-coding regions such as enhancers to achieve dramatic trait effects – despite such regions dominating GWAS signals overall^11^. Because it seems likely that non-coding mutations *are* able to strongly disrupt the binding of individual transcription factors, this might imply that (except in 5’ UTR and 3’ UTR regions) no one transcription factor plays an essential role in the overwhelming majority of cases. Nonetheless, we still observed several cases implicating only non-genic sites, for example an intronic signal for decreasing height (rs570873498;AF=0.0002) at SLC12A1, a gene known to be associated with height and Bartter syndrome, whose symptoms include growth retardation^12^. We anticipate that despite their modest effect sizes and limiting power at present (likely, even if genomes are fully sequenced), the number of non-coding associations will likely increase rapidly in future, once sample sizes become larger. Moreover, our results imply imputation will be highly effective in identifying such associations, even for rare variants.

One unexpected finding for height from our analysis was a cluster of five independent low-frequency associations with height on chromosome 6 (**Supplementary Table 6; Extended data table**), including the rare missense variant rs957675208, in a region not reported by the previous exome sequencing^8^ and exome imputation^9^ analyses, or by HRC-UKB (low imputation INFO). Strikingly, rs957675208 in HMGA1 shows the strongest height-increasing impact of any SNP in the whole dataset, equivalent to gaining 3.5 cm of height. On further examination, three of these four variants are missense mutations and the remaining two 5’ UTR variants are in a gene not annotated in the exome studies. This gives one example of how the complete genome-wide coverage of the GEL-UKB data allows for additional findings compared to previous approaches.

## Methods

### Genomics England high coverage sequencing data

The Genomics England 100,000 Genomes Project was launched in 2013, focusing on rare diseases and cancer. Over 120,000 genomes have been sequenced. It comprises genomes from 73,700 rare disease (disorders affecting ≤1 in 2000 persons) patients and their close relatives, and 46,539 genomes from cancer patients^1^. The GEL reference panel described in this paper is built on the aggregated dataset (aggV2), comprising 78,195 samples from both rare disease and cancer germline genomes. Samples were sequenced with 150bp paired-end reads on the IlluminaHiSeq X platform and processed with the Illumina North Star Version 4 Whole Genome Sequenced Workflow (iSAAC Aligner v03.16.02.19 and Starling small variant caller v2.4.7), and aligned to the GRCh38 human reference genome. The individual gVCF files were aggregated into multi-sample VCF files using Illumina gVCF genotyper and normalised with vt v0.57721. The aggregated multi-sample VCF dataset (aggV2) comprises over 722 million initial called SNPs and short indels (<=50bp). Multi-allelic variants were decomposed into biallelic variants.The l includes 49,641 samples (63.48%) from individuals self-identifying as White British, 4,100 (5.24%) as “Other White”, 2,885 (3.69%) as Pakistani, 1860 (2.3%) as Black, 1,751 (2.24%) as Indian, and 12,277 samples (15.7%) as “Unknown”. The large White British and relatively large South Asian sample size made GEL an ideal reference panel for phasing and imputing UK Biobank, which has a similar ethnic composition^5^. According to the self-reported data, only 27,346 samples (34.97%) are have no relatives in the reference panel. 11,584 (14.81%), 32,679 (41.79%), and 6,586 (8.43%) samples are one of 2, 3 and >3 family members in the dataset respectively. We identified 12,816 (16.39%) samples as members of duo families and 35,106 (44.9%) as members of trio families, while 30,273 (38.71%) samples are treated unrelated for phasing (**Supplementary Notes**).

### Quality Control

Prior to the quality control (QC) described here, sample level QC was carried out by Genomics England informatics team on variants called one sample at a time. We conducted additional quality control by pooling information across samples, to remove false positive sites. Specifically we utilised aggregated VCFs, considering genotype quality, depth, missingness, allele balance, Mendel errors, Hardy-Weinberg equilibrium, and gnomAD^6^ allele frequency concordance.

Because singletons observed in unrelated samples are very hard to phase accurately these sites were removed. We applied two sets of QC rules. First, we applied a stringent rule set applied to all sites, including those *de novo* in Genomics England and very rare sites. Second, we applied a more lenient group of filters for relatively common sites (AF>0.001) that additionally showed support from independent external datasets (TOPMed, HRC, 1000 Genomes, GnomAD), to avoid removing a proportion of genuine sites (e.g. for a modest number of Mendel errors). For these sites, if they failed our stringent filters but passed with somewhat less stringent missingness, Mendel error and gnomAD frequency concordance thresholds, we included them, after separate phasing conditional on the phase of sites passing the more stringent thresholds, i.e. in a manner which did not impact the stringent sites. These sites were incorporated in the final dataset, but with a QC flag indicating their slightly lower reliability. Overall, our filters reduced the initial number of sites from 722 million to 342 million. (**Supplementary Notes and Supplementary Table 1**)

### Phasing the GEL reference panel

We used a multi-stage phasing strategy leveraging the relatedness within GEL, in particular allowing phasing of singletons were possible.

1. We used the makeScaffold software (https://github.com/odelaneau/makeScaffold) to determine the phase of duo and trio samples (**Supplementary Notes**) by direct transmission information (this phases most sites in these samples).
2. For remaining unphased genotypes in these related samples, with phases undetermined due to heterozygosity or missing data, phases were inferred using SHAPEIT4.2.2^13^, with the phased genotypes from step 1 as a scaffold.
3. To phase genotypes in the unrelated samples, we first phased the common variants (AF > 0.01) one chromosome at a time, using SHAPEIT4.2.2 and now using the genotypes (at these common sites) from step 1 and 2 in the related samples as a reference panel.
4. Finally, to phase the remaining sites: genotypes at rare variants in unrelated samples, we using SHAPEIT4.2.2 with the phased samples from steps 1-2 as a reference panel, and the phased common variants from step 3 as a scaffold for these samples.
5. For sites only passing our lenient filters (see “**Quality Control**” section above and **Supplementary Notes**) we used the results of step 4, for the sites on the UKB Axiom array sites passing the stringent filters, as a scaffold, and then used SHAPEIT4.2.2 on the remaining genotypes.

Phasing for steps 1 and 3 was done at the entire chromosome level; for steps 2 and 4 was carried out in regions of approximately 300,000 sites, with 30,000 sites on each side as buffer. The resulting phased regional segments were merged and concatenated using bcftools^14^. These phasing steps were computationally intensive, and took about 6,500 CPU days in total to accomplish. The phased reference panel is stored in VCF format and has been made available for all Genomics England registered users on the GEL trusted research environment.

### Estimation of 1000 Genome trio phasing switch error rate

Phasing accuracy is important for direct biological interpretation of variants within GEL, as well as ensuring high-quality imputation in other samples and other downstream applications. We assessed the ability of the GEL panel to phase such external samples. Specifically, we phased the parents of mother-father-child trios included in the 1000 Genomes Project (but not HRC or GEL) using the reference panels from HRC and GEL. We then assessed the resulting phase accuracy, by comparing phased haplotypes to those directly inferred using inheritance patterns to the child in each trio. The HRC reference panel was lifted over from the GRCh37 to the GRCh38 reference genome using GATK Picard LiftoverVCF^15^. The original GRCh37 HRC reference panel has 39,131,578 autosomal variants. 13,813 variants were removed either due to the incompatibility between reference genomes or mismatching chromosome between the two reference genomes. The resulting autosomal GRCh38 HRC reference panel contain 39,115,765 variants and 27,165 samples. 1000 Genome samples within the HRC reference panel were removed.

We analysed only sites passing 1000 Genome data^16^ filters. The phasing test was carried out on 589 trio families from diverse ethnic backgrounds, using SHAPEIT 4.2.2^13^. We tested all the heterozygous 1000G sites for each individual reference panel, yielding a total of 1.04 × 10^9^ heterozygous sites (1.76 million per trio family) for the HRC panel and 1.16 × 10^9^ (1.9 million per trio family) for the GEL panel.

### Imputation testing of 1000 Genomes samples

We used 2,405 1000 Genomes samples to test the relative performance of imputation based on the GEL, TOPMed and HRC imputation panels. We first performed quality control on the 1000 Genomes data, by removing sites which either possess a missingness larger than 5% or failed a Hardy Weinberg equilibrium test, by having a p-value smaller than 10^−10^ in any of the 26 1000 Genome populations. We then masked genotypes in 1000 Genomes sequencing samples, except the sites existing in the UK Biobank Axiom array, to mimic imputation using this array. This gave 716,473 bi-allelic SNPs across all autosomes. The pseudo-SNP array dataset was then phased one chromosome at a time using SHAPEIT4.1.2^13^. TOPMed imputation was carried out using the TOPMed imputation server with the TOPMed r2 reference panel and the imputation software minimac4 1.5.7^17^. IMPUTE5^18^ was used to impute from the GEL and HRC reference panels. We stratified imputation results into 6 groups : 661 African (AFR), 347 American (AMR), 504 Eastern Asian (EAS), 489 South Asian (SAS), 313 non-Finnish European (NFE) samples and 91 British (GBR) samples.

### UK Biobank imputation using the GEL reference panel

The UK Biobank SNP array data consists of 784,256 autosomal variants. We removed the set of 113,515 sites identified by the previous centralized UK Biobank analysis as failing quality control^5^ and an additional set of 39,165 sites failing a test of Hardy-Weinberg equilibrium on 409,703 White British samples, with the p-value threshold of 10^−10^. The resulting UK Biobank SNP array data was mapped from the GRCh37 to GRCh38 genome build, using the GATK Picard LiftOver tool. Alleles with mismatching strand but matching alleles were flipped. 495 sites were removed due to incompatibility between the two reference genomes, resulting in a final SNP array incorporating 631,081 autosomal variants that we used for phasing and imputation. Haplotype estimation of the SNP array data is a prerequisite for imputation. Phasing was carried out one chromosome at a time using SHAPEIT4.2.2 without a reference panel, using the full set of UK Biobank samples. We ran SHAPEIT4 using its default 15 MCMC iterations and 30 threads. The runtime varied from 2 hours to 30 hours for each chromosome. Imputation of normal filter set and lenient filter set SNPs was carried out independently. Autosomal imputation using the GEL reference panel was performed using IMPUTE5 (v1.1.4). The SNP array data was divided into 408 consecutive and overlapping chunks with roughly 5mb for each chunk and 2.5mb buffer across the genome, using the Chunker program in IMPUTE5^18^ and each chunk was further divided into 24 sample batches with each batch containing 20,349 samples. IMPUTE5 was run on each of the 9,792 subsets using a single thread and default settings, at a speed less than 4 minutes per genome, resulting in a total time of around 1,200 CPU days to impute all UK Biobank samples.

### Genome-wide association studies

We selected four quantitative traits to demonstrate the GWAS performance of the GEL imputed UK Biobank data (GEL-UKB), compared to the HRCUK10K imputed UKB (HRC-UKB) data on 429,460 white British samples. These traits are standing height (HEIGHT), body mass index (BMI), systolic (SBP) and diastolic (DBP) blood pressure. Variants with minor allele count lower than 5 are not included in testing. The trait measures are transformed using rank inverse normal transformation (RINT) within sexes to ensure normally distributed input phenotypes and reduce the likelihood of false positives due to outliers.

Samples between 40 to 70 years old are included and for each data point, outliers that are above±4 standard deviation from the mean value were removed^5^. SBP and DBP values are based on automated blood pressure readings, substituting in manual reading values when automated readings are not available. We calculated the mean SBP and DBP values from two automated (n = 418,755) or two manual (n = 25,888) blood pressure measurements. For individuals with one manual and one automated blood pressure measurement (n = 13,521), we used the mean of these two values. For individuals with only one available blood pressure measurement (n = 413), we used this single value. After calculating blood pressure values, we adjusted for blood pressure-lowering medication (n=94,289) use by adding 15 and 10 mmHg to SBP and DBP, respectively^19^, for individuals on such medication.

GWAS effect size estimates and p-values were obtained using REGENIE^7^.. We used the UKB SNP array data to estimate the LOCO predictors in REGENIE Step 1 and the imputed data for Step 2, accounting for sex, age, sex squared, sex × age, and 20 principal components as covariates^7^. The association tests for GEL imputed UKB (GEL-UKB) and HRCUK10K imputed UKB (HRC-UKB) used the identical setup. The HRC-UKB summary statistics of the association tests were mapped using Picard LiftOver from GRCh37 to GRCh38 to compare the results with GEL-UKB. In all analysis, we used an INFO threshold of 0.3 for common imputed variants (MAF>0.05) and 0.8 for rare imputed variants (MAF≤0.05). **Supplementary Figure 5** shows higher INFO threshold are effective for detecting false positive rare associations.

### Bayesian fine-mapping

Bayesian fine-mapping credible set size comparison was carried out on 1,660, 711, 505 and 546 non-overlapping regions for HEIGHT, BMI, SBP and DBP respectively based on HRC-UKB GWAS summary statistics. These regions were defined by the following procedure. First, candidate regions were identified with width 0.125 centiMorgans plus 25 kb on each side of a significant marker. Overlapping candidate regions were successively merged until there are no remaining regions overlapping. We removed 60, 30, 33, and 51 regions for above traits respectively, in which GEL-UKB showed no significant sites (p-value < 5 × 10^−8^ in GWAS) for each trait. The recombination rate is based on the HapMap genetic map^20^. The detail description of this approach can be found in Maller et al., and Bycroft et al.^5,21^

For each region, we assume a single causal variant – call this model *M*. Given this, define model *M*_*i*_ to be the model where SNP *i* is the causal variant. We seek the probability of *M*_*i*_ given the data and that model *M* is true. This posterior *Pr*(*M*_*i*_|***X***, *M*) can be written in terms of the Bayes factor relating the probability of the data given *M*_*i*_ versus the probability of the data under the null model with no associated SNP in the region, *BF*_*i*_. Further, *BF*_*i*_ can be approximated by an asymptotic Bayesian factor (*ABF*_*i*_):

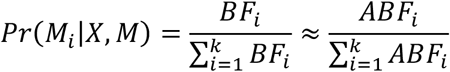

*ABF*_*i*_ can be calculated using the standard error (*V*_*i*_) and Z score (z) estimated by REGENIE^5^. In each region, the smallest possible 95% credible set of potential causal markers can be obtained by successively including the sites with the highest probabilities, to accumulatively reach 0.95. Model *M* requires a prior for the (Gamma distribution) on effect sizes; we choose this prior W to have parameters 0.2^2^ and 0.02^2^, but found the results are not particularly sensitive to the choice of the prior.

### Conditional joint analysis: step-wise regression

A standard GWAS uses marginal model considering one variant at a time, while a joint model considers all the selected variants and estimates their joint effect simultaneously. In order to remove rare variant signals that are explained by stronger signals at more common nearby SNPs^8^. We performed a conditional joint analysis via a stepwise forward selection procedure, considering each chromosome separately. First we defined the set **S** of genome-wide significant variants in one chromosome (P-value < 5 × 10^−8^) in the marginal regression using REGENIE. We initialized a set of variants **R** as the most significant variant in the marginal regression. Given the current value of **R**, we calculate the P-value of all the remaining variants in **S** one at a time, conditioned on **R** and the covariates used for the initial GWAS. We then move the variant with the smallest conditional P-value from **S** to **R**, until this smallest P-value is no longer genome-wide significant. This approach identifies a set of variants that are independently significant, and account for all the genome-wide association signals (note that this set is not unique), while also accounting for linkage disequilibrium between sites. To identify rare causal variants within UKBB found using GEL-UKB imputation, we considered only those variants found by this stepwise forward selection approach. The full conditional joint analysis results can be found in the **Extended data table**.

## Data Availability

All data produced in the present study are available upon reasonable request to the authors

## Data availability

The GEL haplotype reference panel is available within the GEL trusted research environment to approved researchers only. The imputed UK Biobank data imputed using the GEL haplotype reference panel is available to those with approved access to the UK Biobank resource and described on the UK Biobank showcase here https://biobank.ctsu.ox.ac.uk/crystal/field.cgi?id=21008

## Acknowledgements

We thank the Wellcome Trust for funding (200186/Z/15/Z to JM, SM) and (212284/Z/18/Z to SM). Work conducted under UKB applications (48031 and 27960). This research was made possible through access to data in the National Genomic Research Library, which is managed by Genomics England Limited (a wholly owned company of the Department of Health and Social Care). The National Genomic Research Library holds data provided by patients and collected by the NHS as part of their care and data collected as part of their participation in research. The National Genomic Research Library is funded by the National Institute for Health Research and NHS England. The Wellcome Trust, Cancer Research UK and the Medical Research Council have also funded research infrastructure. This work is part of the research portfolio of the National Institute for Health and Social Care Research Barts Biomedical Research Centre. MC is funded by the Barts Charity and is an NIHR Senior Investigator alumnus.

