## Supplementary materials for "A Genomics England haplotype reference panel and the imputation of the UK Biobank"

### Supplementary Note 1: GEL reference panel QC details

The GEL reference panel is built on the aggregated dataset (aggV2), comprising 78,195 samples from both rare disease and cancer germline genomes. Samples are sequenced with 150bp paired-end reads on the IlluminaHiSeq X and processed with the Illumina North Star Version 4 Whole Genome Sequenced Workflow (iSAAC Aligner v03.16.02.19 and Starling small variant caller v2.4.7). The resulting gVCF files are aligned to the GRCh38 human reference genome. Then, the individual gVCF files are aggregated into multi-sample VCF files, using Illumina gVCF genotyper and normalized with vt v0.57721. Sample level quality control has been carried out by Genomics England, including sample contamination less than 0.03, ratio of SNP heterozygous to homozygous calls less than 3, total number of SNPs between 3.2M to 4.7M per sample, array concordance greater than 90%, median fragment size greater than 250bp, excess of chimeric reads less than 5%, percentage of mapped reads greater than 60% and the percentage of AT dropout less than 10%. Finally, the resulting aggregated VCF dataset (aggVCF) was created, comprising over 722 million SNPs and short indels (≤ 50bp). Multi-allelic variants were decomposed into bi-allelic variants. The average genome-wide coverage of the aggVCF samples is 42.2 and median mean coverage is 39.11.

The GEL variants are called individually. A small rate of genotyping error per individual may therefore cause many false positive sites. To filter such sites, in addition to the sample level QC carried out by Genomics England we applied further site level quality control based on the aggregated VCFs:

- **Genotype quality (GQ) + depth (DP)** : Individual genotypes with either GQ < 15 or DP <10 were marked as missing.
- **Missingness**: we removed sites with missing rate higher than 5%, including missingness induced by the GQ + DP filter.
- **Allele balance (ABhet):** allele depths (AD) for REF and ALT are expected not to have a huge discrepancy for each heterozygous individual genotype. For each site, we first calculated the allele balance for each heterozygous call at that site, i.e. AD_REF/(AD_REF + AD_ALT). We then marked cases where 0.25 < ABhet < 0.75 as "pass" in that individual. Sites with a pass rate below 75% were removed.
- **Mendel**: We removed sites with more than 3 Mendelian errors among all duo and trio families for sites with allele frequency < 0.001, or 7 Mendelian errors for sites with allele frequency >= 0.001.
- **Hardy-Weinberg equilibrium (HWE):** Sites where the Hardy-Weinberg Equilibrium (HWE) p-value in self-reported White British samples $< 10^{-5}$ were removed.
- **gnomAD allele frequency (gnomAD)**: We removed sites that showed a discrepancy in allele frequency between GEL and gnomAD. To do this we used a Fisher's exact test for allele frequency difference and a p-value threshold of $10^{-10}$.

- **Unrelated singletons**: we removed singletons that did not occur in related families.
- **Lenient filter setting for common variants**: we chose a set of more lenient filters for those relatively common sites (AF>0.001) found in at least one of the external datasets (TOPMed[1], HRC[2], 1000 Genomes[3], GnomAD[4]). For these sites we used a missingness threshold of 25%, a Mendel error threshold of 250 per site and gnomAD allele frequency filter p-values of $10^{-20}$. All other filters on GQ, DP, ABHet and HWE were kept as above. We generated a file flagging sites retained/recovered by this filter but failing our more stringent QC rules.

A break-down quantifying the sites removed by each successive filter is shown in **Supplementary Table 1**. The final reference panel has 342,560,554 autosomal variants. The overall Ts/Tv ratio increased from 1.1 to 1.8 after filtering.

#### Sample relatedness

The sample relatedness in the reference panel is high. According to the self-reported data, only 27,346 samples (34.97%) found no other relatives in the dataset, whereas 11,584 (14.81%), 32,679 (41.79%), and 6,586 (8.43%) samples 1, 2 and >2 first degree relatives in the dataset, respectively. Among the related samples, 17,871 (22.85%) are marked as proband, 15,908 (20.34%) as mother to the proband, 12,409 (15.8%) as father to the proband, 3,149 (4.03%) as siblings to the proband, and 1,512 (1.93%) as other relatedness, such as grandparents or cousin to the proband. High relatedness improves the performance of Mendel error filtering and phasing accuracy, for example allowing phasing of singletons by transmission: singletons cannot otherwise be phased by the phasing algorithms. We estimated that the reference panel contains approximately 63,000 unique genomes (a 20% reduction in sample size).

To identify parent-child relationship for phasing, we combined information from self-reported relatedness, IBD (identity by descent) and Mendel errors calculated using PLINK[4] . Firstly, 30,000 autosomal variants that meet the following criteria are randomly selected for the analysis: (1) passing the mean genotype quality and depth filter; (2) passing allele balance filter; (3) missingness < 1%; (4) inbreeding coefficient > -0.1; (5) LD-pruned r2 < 0.1 with window size of 500Kb; (6) the Hardy Weinberg equilibrium test p-value > 0.01; (7) in the set of 1000 Genomes phase 3 data; (8) excluding high LD sites identified in Price et al, 2008[5]. We then carried out the following procedure on the selected variants. We selected samples with pairwise IBD0 < 0.1 and IBD1 > 0.7 as potential parent/child pairs. For all potential parent/child pairs matching the self-reported relationships, we identified Mendel errors, separating duo (parent-child) and trio (mother-father-child) families. Where IQR is the inter-quartile range and Q3 is the upper quartile value, the Mendel error cut-offs are Q3+1.5IQR, and Q3+4.5IQR in terms of their mean for trios and duos in order to identify mislabelled and uniparental disomy (UDP) cases. There are 54 potentially UDP cases, often occurring on one chromosome in an affected child-parent pair, and given these small numbers we simply removed all UDP affected pairs and treated them as unrelated. Furthermore, we marked samples as unrelated when self-reported parenthood was inconsistent with the self-reported age, requiring that the parent should be at least 14 years older than the child. Through this procedure, we identified 12,816 (16.39%) samples as in a duo family and 35,106 (44.9%) in a trio family. 30,273 (38.71%) samples were treated as unrelated for phasing.

#### Supplementary Note 2: GEL common variant associations

GEL imputed UKB (GEL-UKB) has 0.6 million fewer common variants (AF>0.01) than HRC-UKB[5]. Common variants constitute the bulk of GWAS findings, since higher allele frequencies always yield higher power. In this note, we will argue that this shortage of common variants compared to HRC-UKB is most likely due to false calls in HRC-UKB.

The GEL reference panel contains 9.2 million common variants (AF > 0.01), compared to 12.8, 9.7, and 9.8 million within the TOPMed, HRC, UK10K and UKB datasets respectively (**Supplementary Table 2**). Over 1 million UK10K common variants (more than 10% of the total UK10K common variant calls) are not found in either GEL or TOPMed. Given that the UK10K is a much smaller reference panel in terms of sample size, and lower coverage (7x) compared to GEL and TOPMed, this suggests an inflation of the number of common variant calls in the UK10K reference panel. These variants are present in the HRC-UKB imputation dataset, because this used both the HRC and UK10K reference panels.

**Supplementary Table 2** also shows that GEL has over 3 million fewer common variants than TOPMed. However, almost all (98.4%) of these TOPMed common variants are found in GEL, but simply at lower allele frequencies. Similarly, TOPMed has 4.8 million more common variants than HRC, among which 3 million variants were present in HRC, with a lower allele frequency. GEL and HRC consist predominantly of samples with European ancestry, while nearly half of the samples in TOPMed have either African or South American identified ancestry. These differences in sample ethnicity can explain the shifted allele frequencies, but not the discrepancy between GEL and UK10K.

In **Supplementary Figure 6**, we identified those significant associations that are unique to each dataset (GEL-UKB or HRC-UKB), and compared their allele frequency concordance to TOPMed allele frequencies, as an independent arbiter. Firstly, we note that 35% of the HRC-UKB-unique sites are present in TOPMed, while 69% of GEL-UKB-unique sites are found. This suggests that many of the former category might be false positives. For sites that do find TOPMed matches, we observe that GEL-UKB unique GWAS hits match the TOPMed allele frequency better than HRC-UKB unique GWAS hits. Many variants estimated as having a lower frequency by TOPMed (i.e. being rare) have an inflated allele frequency in HRC-UKB. In spite of having lower concordance in general, the allele frequency of HRC-UKB GWAS hits matched the TOPMed allele frequency quite well, indicating most of the HRC-UKB unique associations, if occurring in TOPMed, are likely to be real.

We further compared the allele frequency concordance between the most highly discrepant variants, with a genome-wide significant P-value ($< 5 \times 10^{-8}$) in one dataset, and a non-significant P-value ($> 5 \times 10^{-5}$) in the other (**Supplementary Figures 7-8**). For those sites passing GEL filters, GEL-UKB shows good allele frequency agreement with TOPMed, while HRC-UKB shows much poorer agreement, even for those sites more significant in the latter case (**Supplementary Figure 8**). For the smaller number of sites passing only the lenient GEL filters (**Supplementary Note 1**), the GEL-UKB-only sites again show good allele frequency agreement between GEL and TOPMed, but for the HRC-UKB-only site, now the HRC and TOPMed frequencies agree well. This suggests that GEL genotypes are likely most accurate at these discrepant sites in all cases, except those where the lenient filters are required. For these sites it appears GEL calls under-identify true non-reference genotypes (**Supplementary Figure 7**). Hence we suggest those lenient filter sites whose allele frequencies disagree with TOPMed or other data sources need to be used with caution, due to the potential for false negatives (we did not find evidence of false positives).

### Supplementary Tables

| | Number of SNPs left after applying the filter (removed %) | Number of Indels/SVs left after applying the filter (removed %) | Total number of variants after applying the filter (removed %) |
|---|---|---|---|
| **Raw** | 630,967,910 | 91,374,497 | 722,342,407 |
| **+ GQ/DP + missingness** | 428,701,462 (-32%) | 55,702,335 (-39%) | 484,403,797 (-32%) |
| **+ABhet** | 411,285,423 (-3%) | 42,963,226 (-14%) | 454,248,649 (-4%) |
| **+Mendel errors** | 410,854,761 (-0.07%) | 41,905,560 (-1%) | 452,760,321 (-0.2%) |
| **+HWE** | 410,764,722 (-0.01%) | 41,868,797 (-0.04%) | 452,633,515 (-0.01%) |
| **+gnomaAD** | 410,628,878 (-0.02%) | 41,815,306 (-0.05%) | 452,444,184 (-0.02%) |
| **+Singleton** | 309,825,243 (-16%) | 31,639,011 (-11%) | 341,464,254 (-15%) |
| **+Additional filters** | 310,844,262 (+0.16%) | 31,716,292(+0.08%) | 342,560,554(+0.15%) |

**Supplementary Table 1: Variant filtering.** The table shows the effect of each filter applied sequentially from top to bottom in terms of the number of variants (SNPs, Indels/SVs and Total variants) and the percentage removed.

| | Not in overlap with | | | | |
|---|---|---|---|---|---|
| | GEL | TOPMed | HRC | UK10K | HRC-UKB |
| GEL (9,284,802) | 0 | 482,255 | 1,497,927 | 853,110 | 403,068 |
| TOPMed (12,830,650) | 197,955 | 0 | 1,628,650 | 2,663,743 | 4,538,480 |
| HRC (8,021,801) | 85,462 | 96,365 | 0 | 244,174 | 373,528 |
| UK10K (9,769,637) | 1,170,315 | 1,428,019 | 2,099,068 | 0 | 1,255,342 |
| HRC-UKB (9,884,806) | 919,686 | 1,121,825 | 2,079,515 | 1,203,865 | 0 |

**Supplementary Table 2: Common variant overlap across reference panels.** By comparing the row dataset to the column dataset, each cell of this table shows the number of common variants unique to the row dataset (AF>0.01 in the row dataset). For instance, the second row and first column shows the number of TOPMed common variants that cannot be found in the entire GEL dataset is 197,955. The numbers in parentheses are the number of total common variants in each dataset. The GEL variant count in this table includes the sites added by the lenient filters. HRC-UKB is the imputed UK Biobank data by HRC and UK10K[6].

| | p-value | INFO | GEL-UKB | HRC-UKB |
|---|---|---|---|---|
| Common associations | $5 \times 10^{-8}$ | 0.3 | 455,392 | 494,215 |
| Rare associations | $5 \times 10^{-8}$ | 0.8 | 31,699 | 30,711 |
| Common functional | $5 \times 10^{-8}$ | 0.3 | 2,701 (0.59%) | 2,747(0.56%) |
| Rare functional | $5 \times 10^{-8}$ | 0.8 | 511(1.64%) | 473 (1.57%) |
| In common to EWAS | $5 \times 10^{-8}$ | / | 26 (76%) | 22 (65%) |
| In common to EWAS | $2.18 \times 10^{-11}$ | / | 24 (70%) | 19 (56%) |

**Supplementary Table 3: Genome-wide significant association counts across all four traits.** Genome-wide significant association counts across all four traits. Common variants are variants with MAF > 0.05 and rare variants are those with MAF ≤ 0.05, excluding the HLA region in chromosome 6. The number of functionally important variants including high and moderate impact variant effect annotated by VEP are presented in the table, followed by the percentage genome-wide significant ($p < 5 \times 10^{-8}$) in the same category. We also showed the associations that are in common with UK Biobank EWAS results and its proportion of all the EWAS finding[7].

| | | GEL-UKB credible set | Overlap | HRC-UKB credible set |
|---|---|---|---|---|
| Height | Shared | 35,665 | 30,308 | 34,872 |
| | Not shared | 557 | 0 | 2,124 |
| | Total | 36,222 | 30,308 | 36,996 |
| BMI | Shared | 25,477 | 22,208 | 26,166 |
| | Not shared | 302 | 0 | 1,714 |
| | Total | 25,779 | 22,208 | 27,880 |
| SBP | Shared | 19,604 | 17,267 | 20,214 |
| | Not shared | 248 | 0 | 1,457 |
| | Total | 19,852 | 17,267 | 21,671 |
| DBP | Shared | 17,732 | 15,693 | 18,572 |
| | Not shared | 274 | 0 | 1,284 |
| | Total | 18,006 | 15,693 | 19,856 |

**Supplementary Table 4: Fine-mapping credible set sizes stratified by the overlapping of GEL-UKB and HRC-UKB credible sets.** Only regions with both GEL-UKB and HRC-UKB credible sizes smaller than 300 are counted. "Shared" rows indicate the number of sites that are shared between GEL-UKB and HRC-UKB datasets and "Not shared" rows show sites that are unique to each.

| Consequences | HRC-UKB* | GEL-UKB* | HRC-UKB** | GEL-UKB** |
|---|---|---|---|---|
| Splice acceptor | 7593 | 41532 | 5637 | 25777 |
| Splice donor | 10730 | 55730 | 8221 | 35493 |
| Stop gained | 16525 | 78227 | 11910 | 50691 |
| Frameshift | 5536 | 102695 | 5000 | 62333 |
| Stop lost | 1510 | 6900 | 1173 | 4366 |
| Start lost | 2406 | 10541 | 1803 | 6730 |
| Missense | 704682 | 2485273 | 526700 | 1673850 |

**Supplementary Table 5: The number of UK Biobank imputed variants broken down by function.** The variant functions are predicted by VEP (release 105). * indicates all variants in the imputed UKB data. ** is the subset of variants with INFO > 0.3 and minor allele count > 5, which are common thresholds for GWAS.

### Supplementary Figures

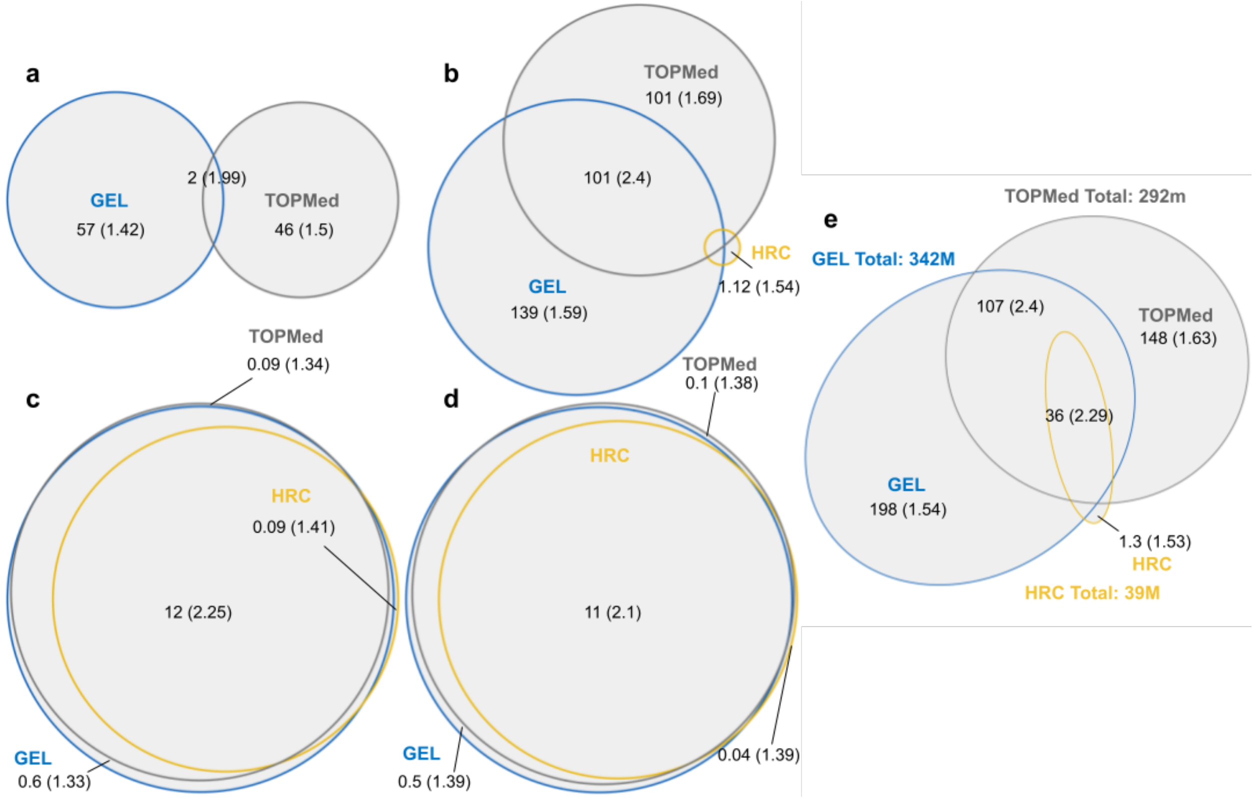


**Supplementary Figure 1: Venn diagram comparing variants from GEL, HRC and TOPMed reference panels.** Allele frequency for variants existing in more than one reference panel is assigned to the highest allele frequency among all the panels. The Venn diagrams show variants with (**a**) $AF < 10^{-5}$, (**b**) $10^{-5} \leq AF < 10^{-4}$, (**c**) $10^{-4} \leq AF < 10^{-2}$, (**d**) $10^{-2} \leq AF < 1$, and (**e**) all variants. The numbers show the variant counts in each region (in millions of variants) followed by the Ts/Tv ratio of these variants.

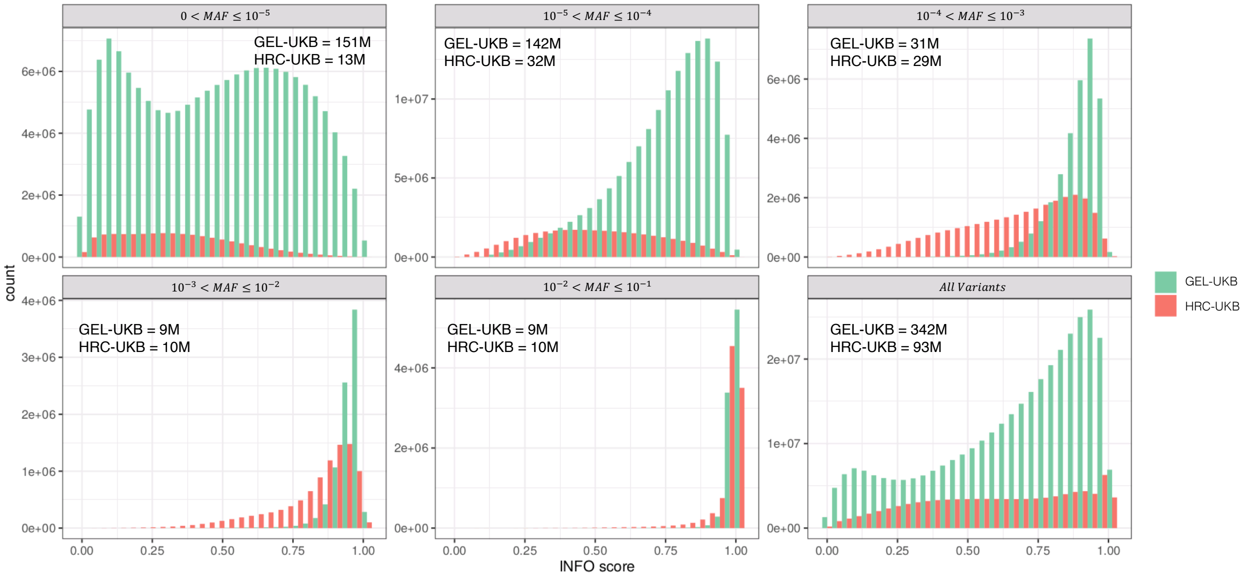


**Supplementary Figure 2: Imputation INFO score histogram comparison between GEL-UKB and HRC-UKB.** Each panel shows the distribution of INFO scores for GEL and HRCUK10K imputed variants in different MAF bins. The total number of variants in each bin is provided in the panel legend.

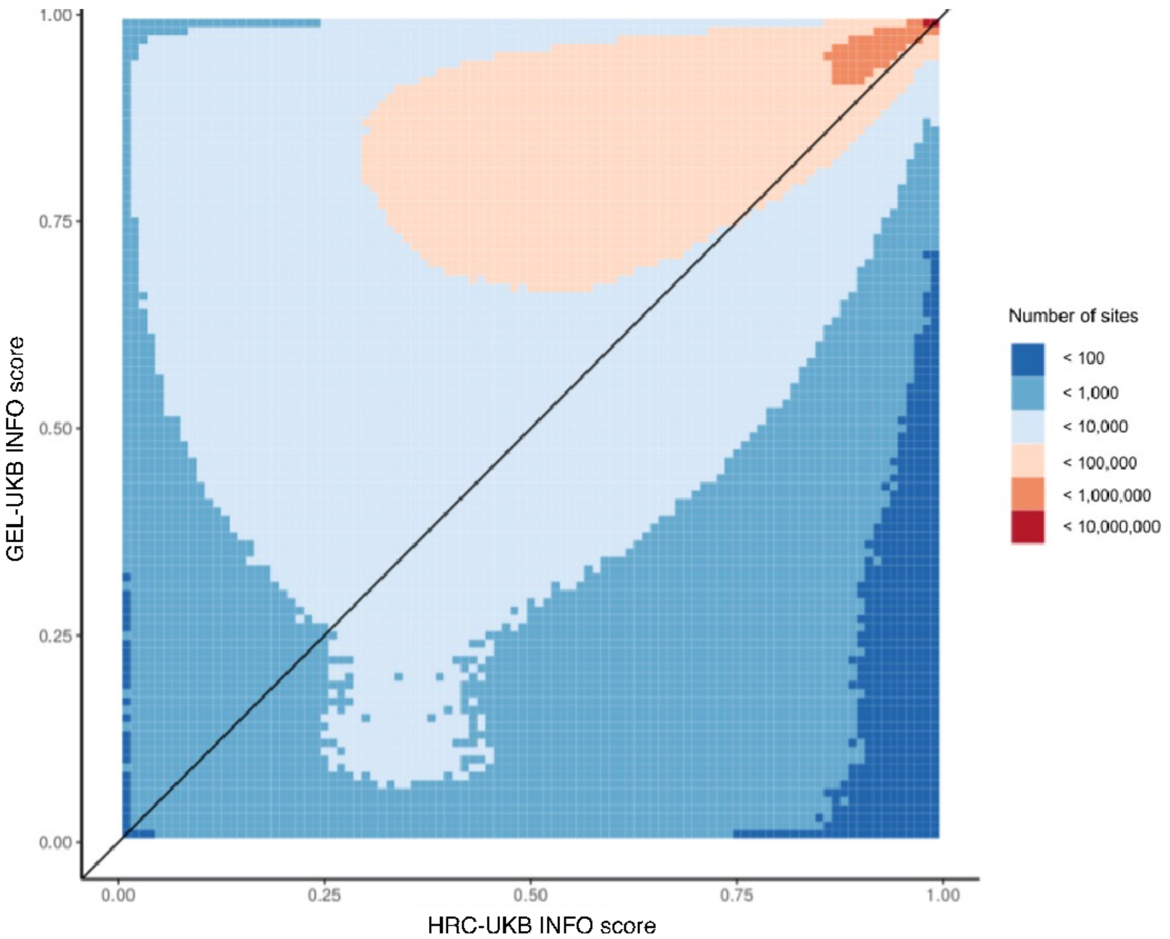


**Supplementary Figure 3: A heatmap of imputed UK Biobank INFO scores from the 65 million sites present in both GEL-UKB and HRC-UKB.** GEL imputation of the UK Biobank (GEL-UKB) shows improved INFO scores for 87% of existing imputed markers in HRC-UKB. The x-axis shows imputation using HRC-UKB, and the y-axis GEL-UKB.

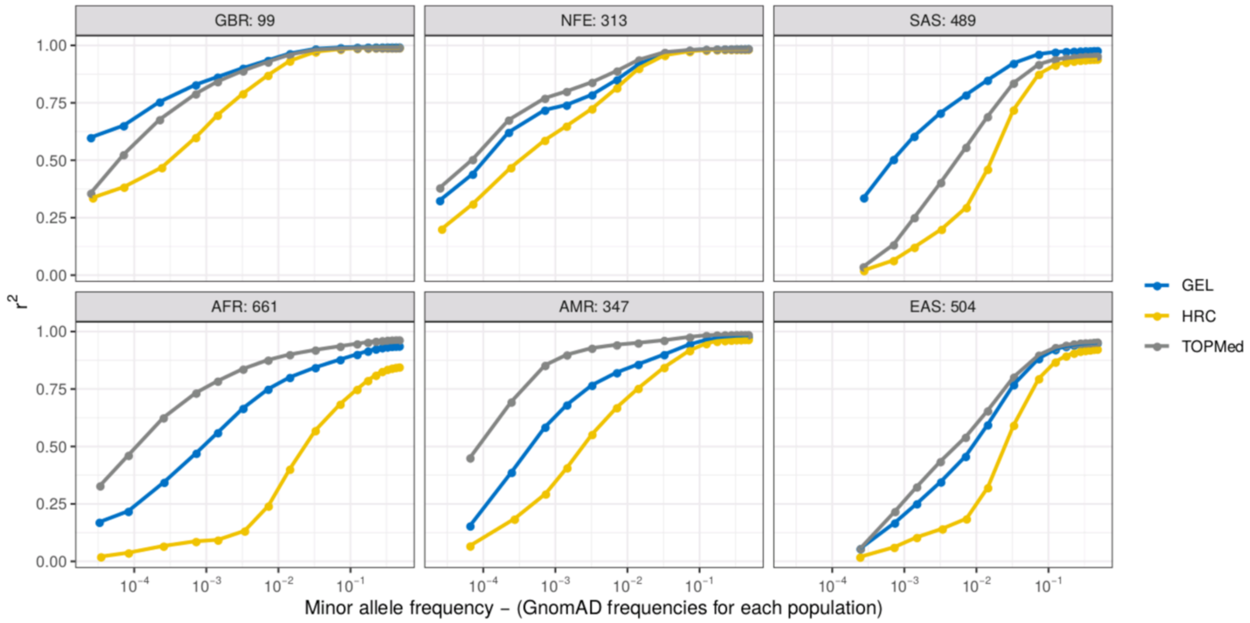


**Supplementary Figure 4: Comparison of imputation performance in 1,000 Genome samples using different reference panels.** The x-axis shows non-reference allele frequency on a log scale allowing focusing on rarer variants. The y-axis is imputation performance ($r^2$). The performance of the reference panels HRC (yellow), TOPMed (grey), GEL (blue) are shown as lines in each plot. The variants are stratified by GnomAD allele frequency (v3.3.1) of their corresponding populations, including GBR (White British), NFE (non-Finnish European), SAS (South Asian), AFR (African), AMR (American), and EAS (East Asian) totalling 2,405 samples. The sample size for each population follows the population labels. The variants are stratified by GnomAD allele frequency (v3.3.1)[4] in the corresponding population.

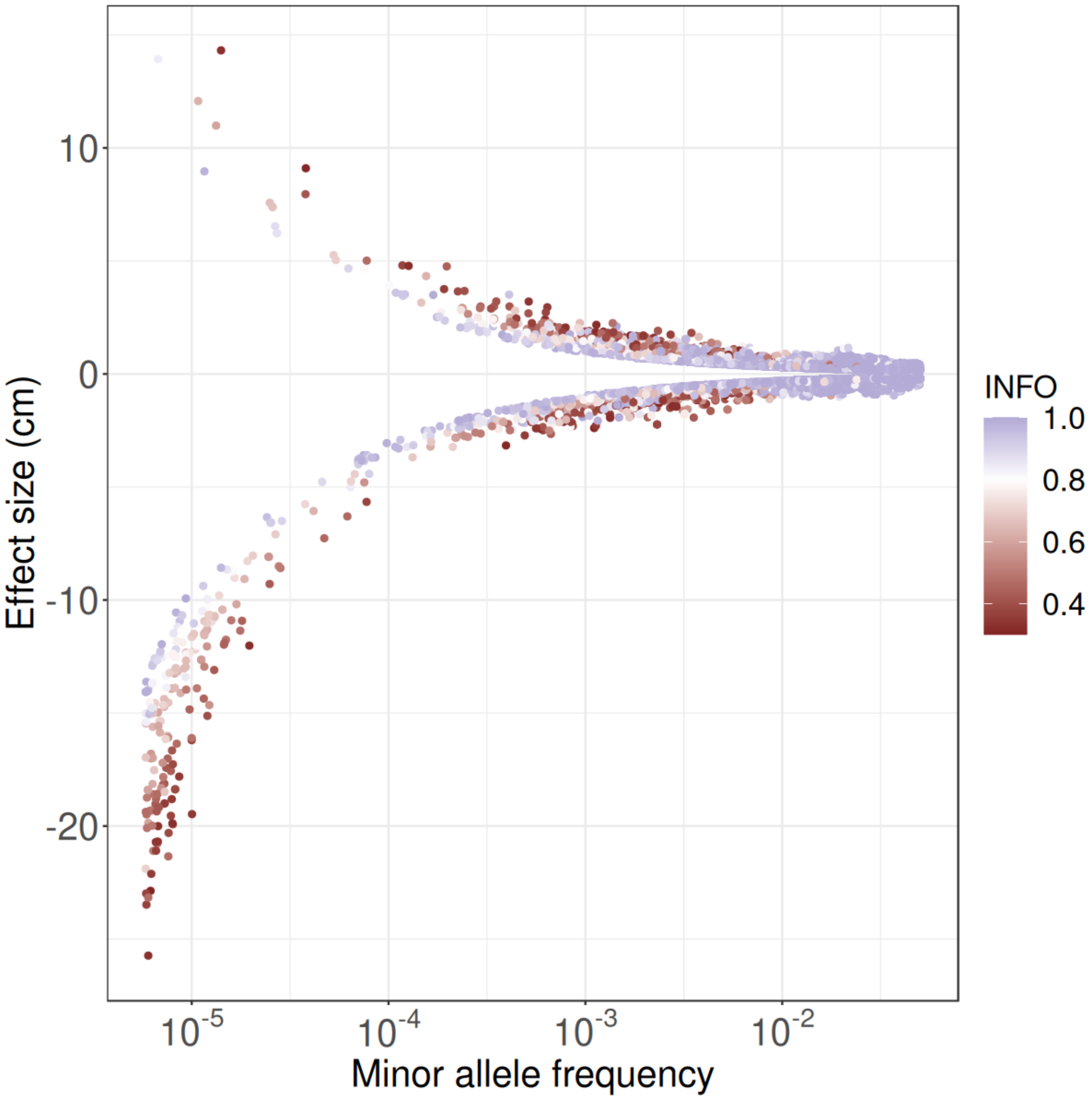


**Supplementary Figure 5: Frequency and effect size plot of rare genome-wide significant associations (MAF < 0.05) for standing height, found using GEL-UKB.** Rare associations require higher effect sizes to be detected. We observe that ultra-rare variants with large effect sizes often possess lower INFO scores. In our GWAS analysis, if not specified, the INFO threshold for rare associations (MAF < 0.001) is 0.8.

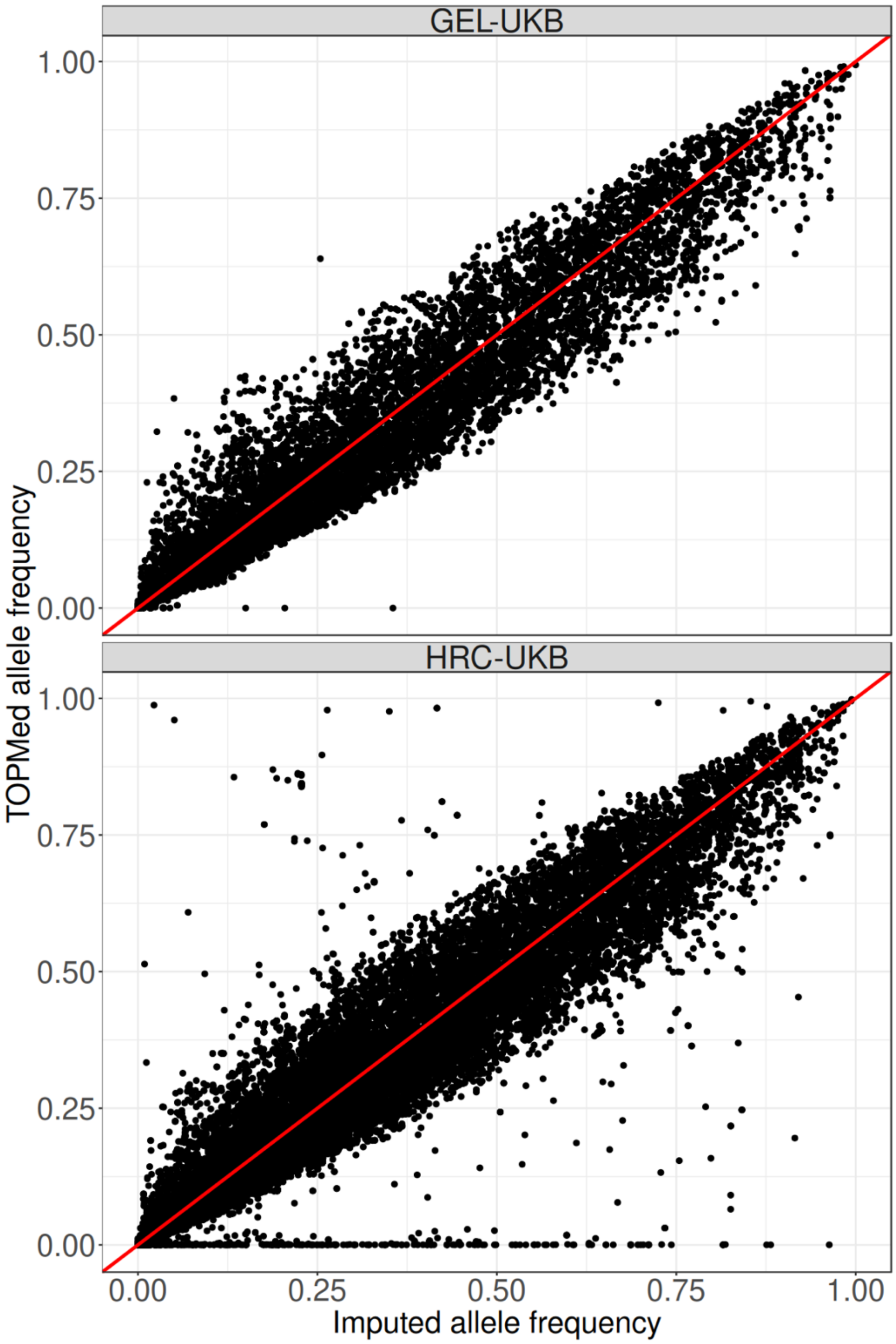


**Supplementary Figure 6: Allele frequency concordance of the GWAS significant sites vs. TOPMed allele frequencies.** For genome-wide significant sites across all four traits, imputed allele frequencies of GEL imputed UK Biobank data (GEL-UKB) and HRCUK10K imputed UK Biobank data (HRC-UKB) (x-axis) are compared to TOPMed allele frequencies (y-axis). Variants within the HLA region or with INFO score below 0.3 are excluded from the plot.

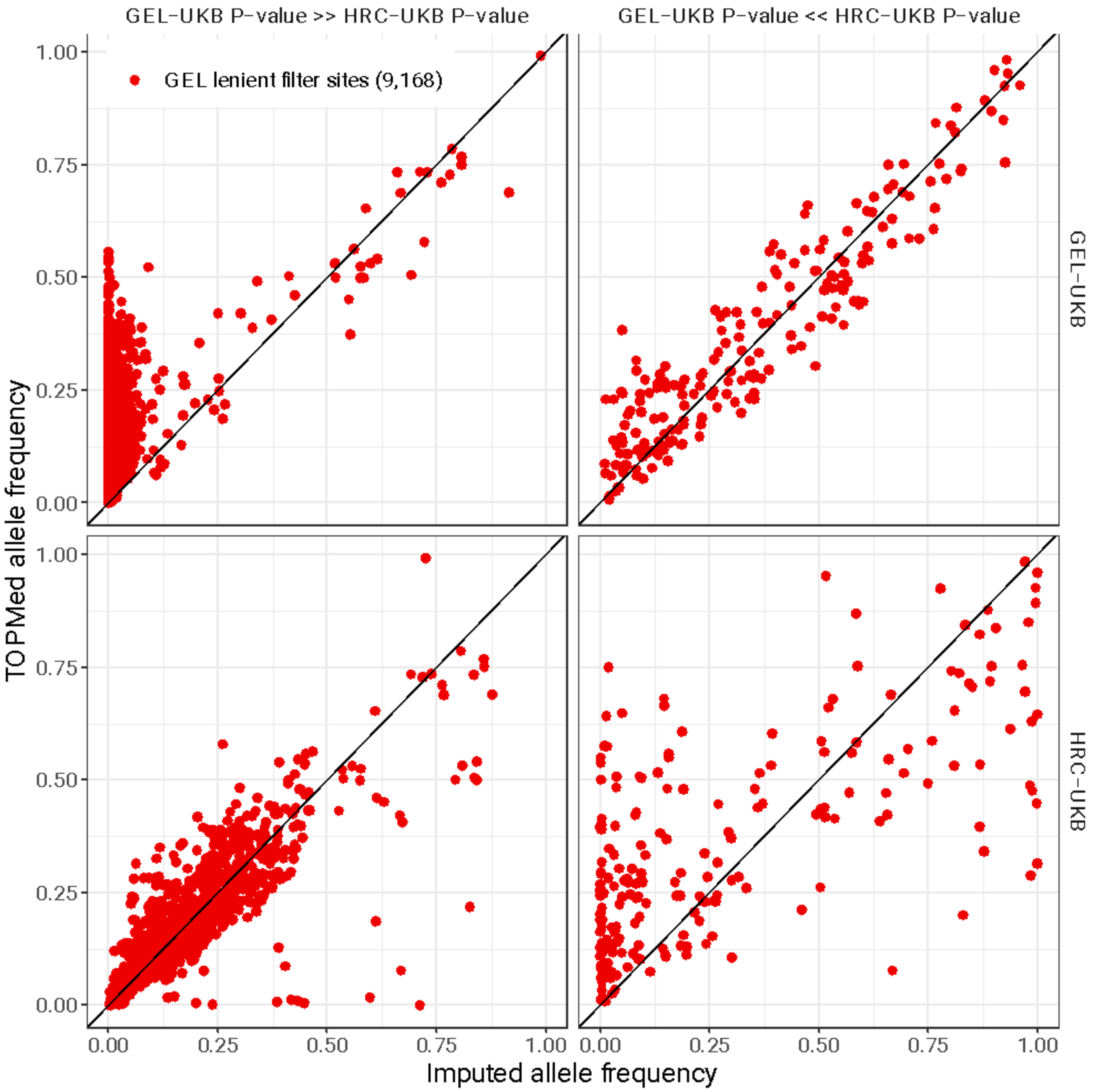


**Supplementary Figure 7: Comparing allele frequency concordance to TOPMed for lenient-filter sites showing large P-value discrepancies between GEL-UKB and HRC-UKB**. Associations that are significant in HRC-UKB but possessing a much bigger P-value ($> 5 \times 10^{-5}$) in GEL-UKB are shown in the left column, for only those sites passing GEL lenient filters but not GEL strict filters. The right column shows the same, but for associations that are significant in GEL-UKB but possessing a much bigger P-value HRC-UKB. The rows compare estimated allele frequencies in GEL-UKB (top row, x-axis) or HRC-UKB (bottom row, x-axis) to TOPMed allele frequencies (y-axes).

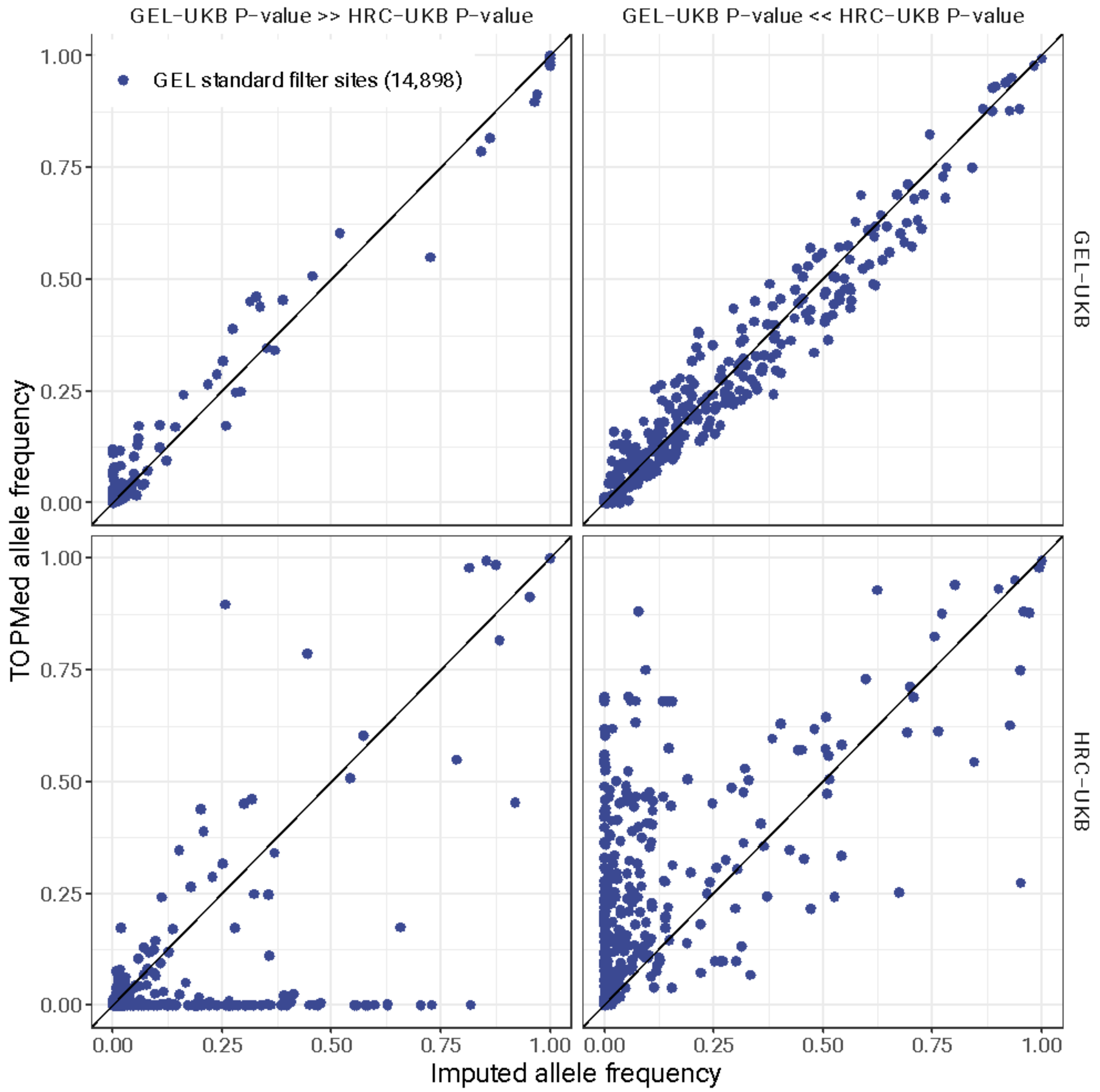


**Supplementary Figure 8: Comparing allele frequency concordance to TOPMed for standard-filter sites showing large P-value discrepancies between GEL-UKB and HRC-UKB**. Associations that are significant in HRC-UKB but possessing a much bigger P-value (> 5 × $10^{-5}$) in GEL-UKB are shown in the left column, for only those sites passing GEL strict filters. The right column shows the same, but for associations that are significant in GEL-UKB but possessing a much bigger P-value HRC-UKB. The rows compare estimated allele frequencies in GEL-UKB (top row, x-axis) or HRC-UKB (bottom row, x-axis) to TOPMed allele frequencies (y-axes).

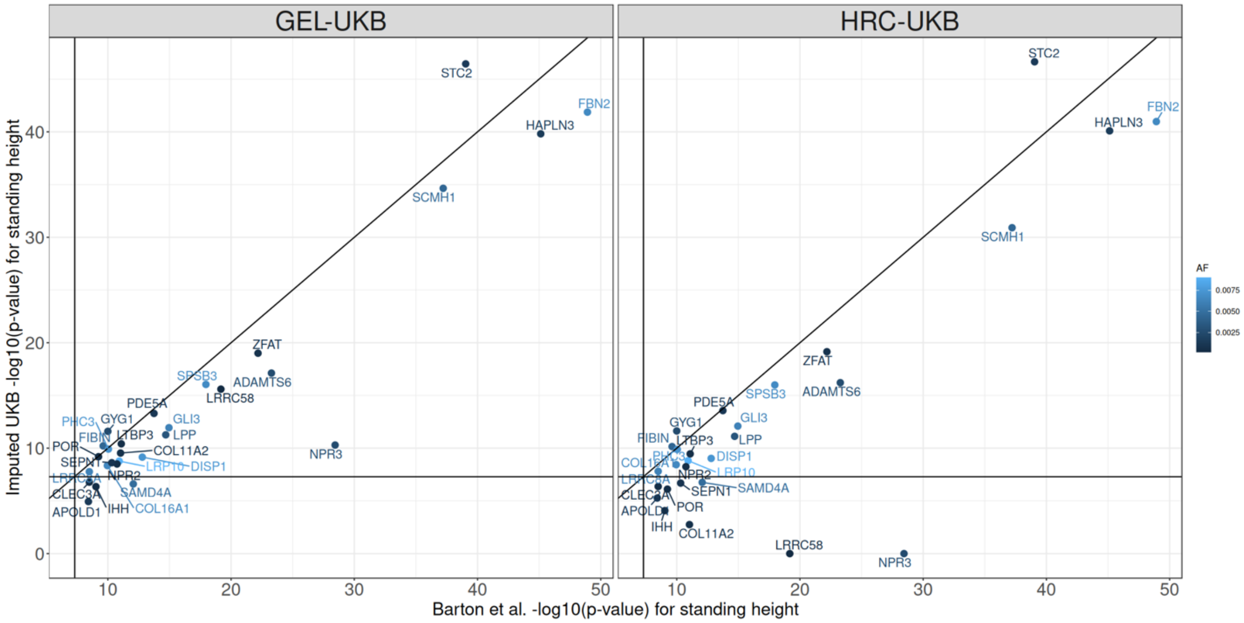


**Supplementary Figure 9: Comparing the p-value of GEL-UKB and HRC-UKB with rare likely-causal coding variants identified by the whole exome imputation[8] (Barton et al., 2021, Table 1).** The x-axis shows the p-value using the exome imputation and the y-axis shows the p-value using GEL-UKB (left) and HRC-UKB (right). The -log p-value is set to be 0, when the variant is not found. The dots are colours according to their allele frequency from the exome imputation data. The horizontal and vertical lines show the genome-wide significant threshold, i.e. $5 \times 10^{-8}$.
